# Analytical performance of a multi-target open real-time PCR assay for simultaneous detection of tuberculosis, non-tuberculous mycobacteria, and drug resistance in a high-burden setting

**DOI:** 10.64898/2026.04.23.26351557

**Authors:** Zeeshan Sidiq, Payal Tyagi, Ankita Anand, Kaushal Kumar Dwivedi, Sanjay Rajpal, Kamal Kishore Chopra

## Abstract

**Background:** Timely diagnosis of tuberculosis and drug resistance remains a cornerstone of effective disease control. Multiplex open molecular platforms capable of simultaneously detecting *Mycobacterium tuberculosis complex* (MTBc), non-tuberculous mycobacteria (NTM), and resistance to first-line anti-tuberculosis drugs could streamline diagnostic pathways.

**Methods:** We conducted a laboratory-based evaluation of two multiplex real-time PCR assays (MTBc/NTM R-Gene® and MTB-RIF/INH R-Gene®) using 300 well-characterized samples, including 150 MTBc-positive culture isolates (including rifampicin-resistant, isoniazid-resistant, and drug-susceptible strains) and 150 MTBc-negative samples (50 NTM isolates and 100 mycobacteria-negative specimens). Composite reference standards included culture, MPT64 antigen testing, and line probe assay corroborated by phenotypic drug susceptibility testing for resistance profiling, with NTM speciation performed using a dedicated line probe assay. DNA extraction was performed using the QIAamp DNA Mini Kit (QIAGEN, Germany), followed by amplification on a real-time PCR platform according to manufacturer instructions. The diagnostic performance was assessed against composite reference standards.

**Results:** The analytical performance for detecting MTBc demonstrated 100% sensitivity and specificity (150/150). NTM detection showed 70·0% sensitivity (35/50) and a specificity of 100%, highlighting limitations in coverage of NTM species. Rifampicin resistance was detected with a sensitivity of 96·0% (48/50) and specificity of 100%, whereas isoniazid resistance detection was 100% sensitive and specific (50/50). Agreement with established reference standards was high (κ=0·76–1·00) within this analytical context.

**Interpretation:** This analytical validation demonstrates that multiplex open real-time PCR assays can accurately and simultaneously detect MTBc, NTM, and rifampicin and isoniazid resistance using culture isolates. While these platforms offer potential advantages in flexibility and expanded resistance profiling, additional studies on clinical diagnostic accuracy, cost-effectiveness analyses, and operational feasibility are required to determine their practical utility and programmatic impact in high-burden settings

## Introduction

Tuberculosis (TB) remains a global public health challenge, with India bearing the highest burden of disease [1]. Rapid, accurate, and scalable diagnostic tools are becoming increasingly important in meeting the TB elimination targets set by the National Tuberculosis Elimination Programme (NTEP) particularly when it comes to early detection of drug-resistant TB. However, traditional diagnostic approaches, such as smear microscopy lack sensitivity, and culture, while regarded as the gold standard, is time-consuming and resource-intensive [2,3,4].

The introduction of nucleic acid amplification tests (NAATs) has substantially improved TB diagnostics, allowing for rapid detection of *Mycobacterium tuberculosis* and rifampicin resistance. Cartridge-based molecular platforms have been widely adopted due to their operational simplicity and faster turnaround time [5,6,7,8]. Nonetheless, these systems are mostly proprietary, haver high per-test costs, and primarily target rifampicin resistance as a surrogate marker for multidrug-resistant TB (MDR-TB). Importantly, these assays have limited ability to detect isoniazid resistance independently, which may result in underdiagnosis of isoniazid mono-resistance and poor treatment recommendations. Furthermore, issues like indeterminate results, reliance on continuous supply chains, and restricted flexibility in assay design continue to hamper their effectiveness in routine programmatic settings [9,10,11].

Another significant diagnostic challenge is distinguishing *Mycobacterium tuberculosis complex* (MTBc) from non-tuberculous mycobacteria (NTM), which is becoming more common in clinical practice [12,13]. Misclassifying of NTM as TB can lead to incorrect treatment and poor patient outcomes. The most widely used rapid molecular assays do not significantly address this issue, highlighting the need for more comprehensive diagnostic tools [14].

Open real-time polymerase chain reaction (RT-PCR) platforms provide a flexible alternative, which enables the simultaneous detection of multiple targets, such as the distinction of MTBc from NTM and the identification of mutations associated with drug resistance. These systems offer potential benefits in terms of cost, flexibility, throughput, and increased diagnostic capabilities including the detection of both isoniazid and rifampicin resistance, by enabling the use of non-proprietary reagents and customized assay designs.

Despite these advantages, there is limited clinical data on the diagnostic performance of open RT-PCR–based assays for the simultaneous detection of TB, NTM, and drug resistance in real-world settings [15, 16]. There is a large knowledge gap about the clinical applicability of multi-target open molecular platforms because majority of the current research has been on closed cartridge or chip-based systems. In contrast to currently used molecular platforms that are largely restricted to single-target detection or rely on proprietary cartridge or chip-based systems, this approach allows for the simultaneous detection of MTBc, differentiation of NTM, and identification of both rifampicin and isoniazid resistance within a single open-platform workflow. This integrated, multidomain capability addresses the two most persistent and clinically important gaps in current diagnostic algorithms, the under detection of isoniazid resistance and limited identification of NTM in routine practice.

In this context, the present study was undertaken to evaluate the analytical accuracy of the Argene® MTBc/NTM and MTB-RIF/INH qualitative real-time PCR assays (bioMérieux, India) for the detection of the *Mycobacterium tuberculosis complex*, differentiation from non-tuberculous mycobacteria, and identification of rifampicin and isoniazid resistance using well-characterized culture isolates. The analytical performance of these assays was assessed using culture and line probe assay as reference standards, with the aim of generating evidence of their potential role as comprehensive molecular diagnostic tools in high-burden settings.

## Methods

### Study design, population, and reference standard

This laboratory-based, comparative analytical validation study was conducted at the New Delhi Tuberculosis Centre, New Delhi-an Intermediate Reference Laboratory (IRL) under the NTEP, India. The study aimed to assess the analytical performance of open real-time polymerase chain reaction (RT-PCR) assays for the detection of MTBc, differentiation from NTM, and detection of drug resistance.

A total of 300 well-characterized culture isolates were included, comprising laboratory available microbiologically confirmed specimens. Samples were stratified into MTBc-positive (n=150) and MTBc-negative (n=150) groups. Although it does not represent the occurrence in clinical settings and may affect predictive values, this 1:1 ratio was deliberately chosen to ensure robust calculations of analytical sensitivity and specificity for each target, a common practice in initial analytical validation studies. The MTBc-positive group consisted of culture-confirmed *Mycobacterium tuberculosis* isolates with predefined drug susceptibility profiles based on GenoType MTBDRplus VER 2.0 Line Probe Assay (Hain Lifescience GmbH, Nehren, Germany), including 50 rifampicin-resistant, 50 isoniazid-resistant, and 50 drug-susceptible isolates. Corresponding phenotypic drug susceptibility testing results obtained using the BACTEC MGIT 960 system were available for these isolates as part of prior routine laboratory characterization. The MTBc-negative group comprised 50 non-tuberculous mycobacteria (NTM) culture isolates and 100 samples negative for mycobacteria, confirmed by culture and/or rapid assays, including MPT64 antigen testing or molecular methods such as Xpert MTB/RIF.

Species identification was established using an MPT64 antigen detection assay, with MPT64-positive isolates classified as MTBc. NTM isolates were identified using the GenoType Mycobacterium CM/AS line probe assay (Hain Lifescience GmbH, Nehren, Germany) for mycobacterial speciation. Drug resistance classification for rifampicin (RIF) and isoniazid (INH) was based on previously established genotypic results obtained using the GenoType MTBDRplus VER 2.0-line probe assay, which served as the primary reference standard for detection of mutations in *rpoB, katG*, and *inhA*. Corresponding phenotypic drug susceptibility testing results available from prior routine laboratory characterization and were used to support resistance classification. A composite reference framework incorporating antigen-based and molecular methods, with supportive phenotypic data, was applied to mitigate the limitations of individual approaches and enhance the robustness of diagnostic classification.

The sample size was selected to enable reliable estimation of diagnostic accuracy parameters. The distribution of samples across resistance profiles and non-tuberculous mycobacteria was structured to ensure adequate representation for subgroup analyses and meaningful performance evaluation.

#### Index tests and procedures

Two multiplex real-time PCR assays (ARGENE®, bioMérieux, India) were evaluated: (1) MTBc/NTM R-Gene® for species identification and (2) MTB-RIF/INH R-Gene® for detection of resistance-associated mutations. Both assays utilize 5⍰-exonuclease (TaqMan) chemistry.

For sample processing, 500 µL of liquid culture was centrifuged at 5000 rpm for 15 minutes, and the pellet was resuspended in 200 µL phosphate-buffered saline (pH 8·0). An internal control (IC2) was added prior to extraction to monitor extraction efficiency and PCR inhibition. DNA extraction was performed using the QIAamp DNA Mini Kit (QIAGEN, Hilden, Germany; Cat. No.: 51304) from 200 µL of processed sample, following the manufacturer’s instructions. DNA was eluted in 50 µL of Buffer AE and used immediately or stored at −20 °C until further analysis.

#### Real-time PCR amplification

PCR reactions were prepared in a dedicated clean area. Each reaction consisted of 15 µL of assay-specific master mix and 10 µL of extracted DNA. Positive and negative controls were included in each run. Amplification was performed on a Bio-Rad CFX96 real-time PCR system using the following cycling conditions: initial denaturation at 95°C for 15 minutes, followed by 42 cycles of 95°C for 10 seconds and 60°C for 40 seconds, with fluorescence acquisition at each elongation/hybridization cycle.

The MTBc/NTM R-Gene® assay targets IS6110 and IS1081 for MTBc detection and the rpoB gene for pan-mycobacterial detection. The MTB-RIF/INH R-Gene® assay detects mutations within the rifampicin resistance-determining region of rpoB (codons 511, 516, 522, 526, and 531), and mutations in katG (including codon 315 and 328) and the inhA promoter region associated with isoniazid resistance. Fluorescence detection was performed across multiple channels in accordance with the manufacturer’s instructions, including FAM (MTBc-specific targets, Rifampicin and INH high level resistance targets), HEX (internal control), ROX (pan-mycobacterial), and Cy5 channel (INH low level resistance targets).

Assay performance characteristics, as defined by the manufacturer, include limits of detection of approximately 15 CFU/mL for MTBc and 0·1 CFU/mL for NTM, and 5 copies/µL for resistance targets. Cycle threshold (Ct) cut-offs of ≤37 cycles for MTBc/NTM detection and ≤42 cycles for resistance detection were applied.

#### Interpretation and analysis

Results were interpreted according to manufacturer-defined criteria. MTBc was defined by detection of MTBc-specific targets, whereas NTM was defined by detection of pan-mycobacterial targets in the absence of MTBc-specific amplification. Resistance to RIF and INH was inferred from detection of mutation-specific signals. Each run was considered valid only if control reactions met predefined criteria. Cycle threshold values were generated using instrument software CFX maestro Version 2.3. Index test results were compared against the composite reference standard, comprising MPT64 antigen testing for MTBc identification, line probe assays for NTM speciation and initial resistance characterization, and phenotypic drug susceptibility testing (MGIT 960) as confirmatory evidence for resistance.

### Ethics approval and compliance

This study was conducted in accordance with the Declaration of Helsinki, with approval from the Institutional Ethics Committee of the New Delhi Tuberculosis Centre, New Delhi (Project/TB/36/392A). Laboratory procedures were performed within an ISO 15189-compliant framework, and reporting adhered to STARD 2015 guidelines and National Tuberculosis Elimination Programme (NTEP), India standards.

## Results

During the study period, 300 culture-confirmed samples were analyzed, including 150 MTBC-positive and 150 MTBc-negative samples. Among the MTBc-positive isolates, 50 were rifampicin-resistant, 50 were isoniazid-resistant, and 50 were drug-susceptible, as determined by the GenoType MTBDRplus VER 2.0 LPA. The MTBC-negative group included 50 non-tuberculous mycobacteria (NTM) isolates and 100 samples with no mycobacteria.

For MTBc detection, the assay identified all 150 MTBc-positive samples, corresponding to a sensitivity and specificity of 100 %. Among the 150 MTBc-negative samples, there were no false-positive results. For NTM detection, the analytical sensitivity was lower at 70%, with 15 isolates not detected, whereas the specificity remained 100%. The limited diversity of NTM species included in this analytical validation may impact the generalizability of this sensitivity. The positive predictive value was 100%, and the negative predictive value was 87%, indicating reliable identification when NTM was present, but some cases could be missed (table1).

**Table 1.** Diagnostic accuracy of the Argene® multiplex real-time PCR assay for MTBc/NTM identification and detection of rifampicin and isoniazid resistance. Note: All performance metrics are derived from analytical validation using culture isolates. The high accuracy and Kappa values reflect strong analytical agreement within a controlled laboratory setting and may not directly translate to clinical performance on patient specimens due to spectrum bias.

| Target | TP / TN /<br>FP / FN | Sensitivity<br>% (95% CI) | Specificity %<br>(95% CI) | PPV %<br>(95% CI) | NPV %<br>(95% CI) | Accuracy %<br>(95% CI) | Kappa |
| --- | --- | --- | --- | --- | --- | --- | --- |
| MTBc detection | 100 / 100<br>/ 0 / 0 | 100 (96.3–<br>100) | 100 (96.3–<br>100) | 100 (96.3–<br>100) | 100 (96.3–<br>100) | 100 (98.1–<br>100) | 1 |
| NTM detection | 35 / 100 /<br>0 / 15 | 70.0 (56.2–<br>80.9) | 100 (96.3–<br>100) | 100 (90.1–<br>100) | 87.0 (79.6–<br>91.9) | 90.0 (84.2–<br>93.8) | 0.76 |
| Rifampicin resistance | 48 / 50 /<br>0 / 2 | 96.0 (86.5–<br>98.9) | 100 (92.9–<br>100) | 100 (92.6–<br>100) | 96.2 (87.0–<br>98.9) | 98.0 (93.0–<br>99.4) | 0.96 |
| Isoniazid resistance | 50 / 50 /<br>0 / 0 | 100 (92.9–<br>100) | 100 (92.9–<br>100) | 100 (92.9–<br>100) | 100 (92.9–<br>100) | 100 (93.0–<br>100) | 1 |

MTBc: Mycobacterium Tuberculosis Complex; NTM: Non-Tuberculous Mycobacteria; True positives; TN: True negatives; FP: False positives; FN: False negatives; PPV: Positive predictive value; NPV: Negative predictive value

For drug resistance detection, the assay correctly identified all 50 isoniazid-resistant isolates, with 100% sensitivity and specificity. Rifampicin resistance was detected in 48 of 50 resistant isolates, giving 96% sensitivity, 100% specificity, and an overall accuracy of 98%. The positive and negative predictive values were high for both resistance targets, supporting confidence in resistance profiling (table1).

Ct value distributions supported assay performance (Figure 1). MTBc targets showed low Ct values (mean 18.9; range 11.3–31.7) well below the cut-off (≤37). Rifampicin resistance (rpoB) yielded higher Ct values (mean 27.7; 21.4–39.9) within threshold (≤42), while isoniazid targets (katG/inhA) showed a narrower distribution (mean 21.7; 17.9–28.1). Ct values were consistently separated from cut-offs, minimizing indeterminate results.

**Figure 1.**
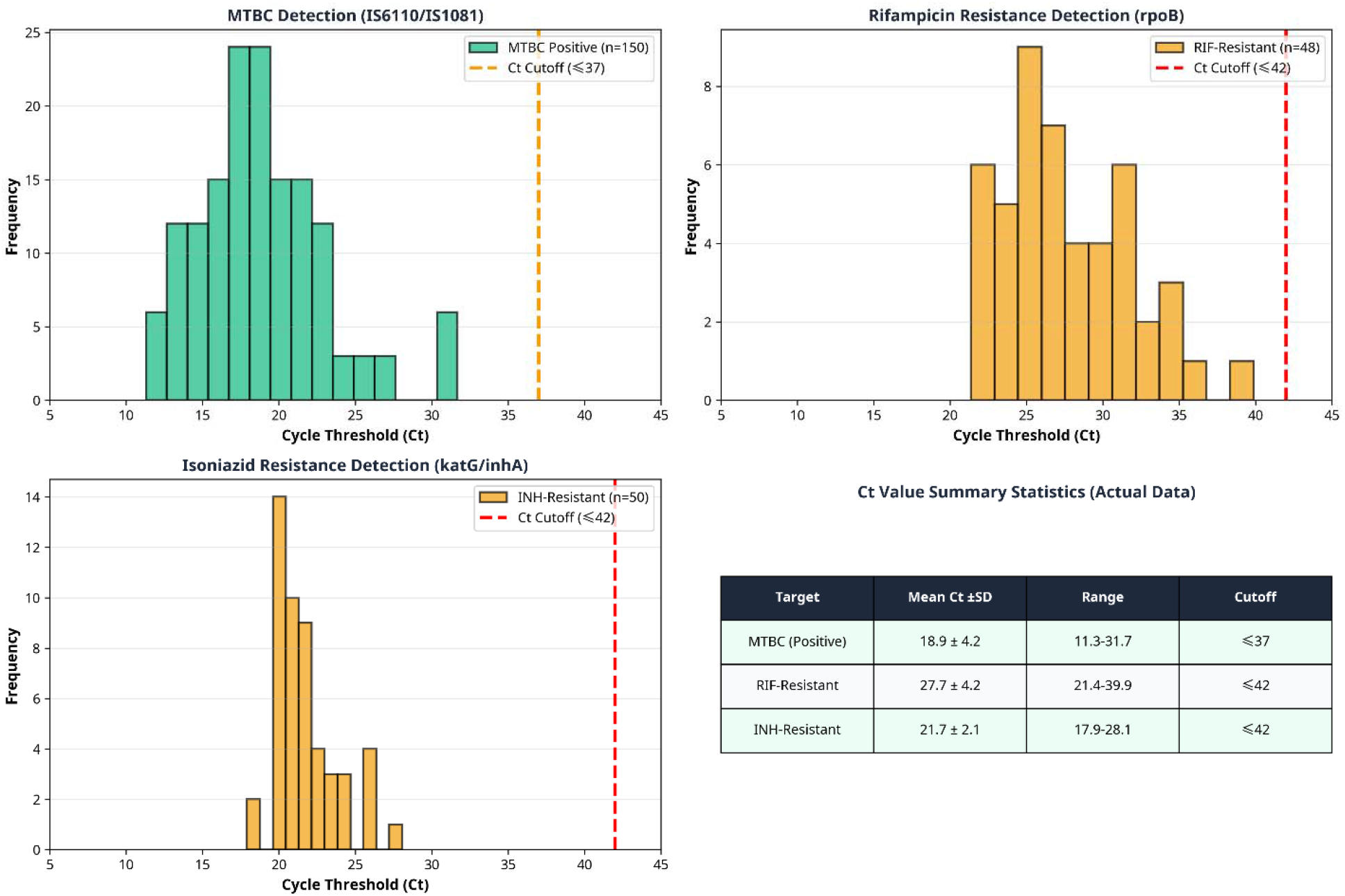
Distribution of cycle threshold (Ct) values for MTBc detection (IS6110/IS1081), rifampicin resistance (rpoB), isoniazid resistance (katG/inhA) targets, and summary statistics of Ct values.

## Discussion

This study demonstrates that a multiplex, open real-time PCR platform can achieve high analytical accuracy for the simultaneous detection of MTBc and key drug resistance markers, while also enabling differentiation from NTM within a single workflow using culture isolates. The high degree of concordance (κ=0·76–1·00), indicating significant to near-perfect analytical agreement with established reference standards, further supports the analytical reliability of the assay in a controlled laboratory setting.

The 100% sensitivity and specificity of the assay for MTBc detection underscore the reliability of nucleic acid amplification tests targeting conserved genomic regions. This performance is comparable to that of widely used molecular platforms, such as Xpert MTB/RIF, which have transformed early TB diagnosis and treatment initiation. In high-burden settings, where diagnostic delays fuel transmission and worsen patient outcomes, such rapid and accurate detection is critical [1, 5,6]. The absence of false positive MTBc results is particularly important, as it minimizes the risk of inappropriate treatment and associated program burden.

The assay demonstrated outstanding agreement with reference standards for drug resistance detection. With just two false-negative results, Rifampicin resistance was detected with 96% sensitivity and 100% specificity. This observation is consistent with the recognized limitations of molecular assays targeting specific rpoB regions, which may miss uncommon or atypical mutations outside of designated hotspots [17,18]. Nevertheless, this performance remains clinically robust given the role of rifampicin as a surrogate marker for multidrug-resistant tuberculosis. Most notably the complete detection of isoniazid resistance (100% sensitivity and specificity), likely due to targeting both katG and inhA, represents a significant advance. This capability addresses a critical gap in rifampicin-focused assays, which often underdiagnose isoniazid mono-resistance, potentially leading to suboptimal treatment regimens [19]. By simultaneously detecting both key first-line drug resistance, this assay enables more precise treatment guidance and prevents the development of MDR-TB.

A key strength of this approach lies in its integrated, multi-target design on an open platform, enabling the concurrent detection of MTBc, differentiation of NTM, and identification of resistance-associated mutations. This is in contrasts to the rapid molecular diagnostics currently in use, which either depend on proprietary cartridge-based systems or are restricted to single target detection. Open-platform assays offer important advantages, including greater flexibility in assay design, independence from closed supply chains, and the possibility for cost optimization through reagent selection and workflow consolidation. These attributes are especially relevant in high-burden countries such as India, where expanding diagnostic capacity under national programs calls for scalable, adaptable, and sustainable solutions. [11,15,20]. The ability to customize assay panels represents a significant advantage for resource-limited settings seeking to strengthen diagnostic infrastructure.

The observed 70% analytical sensitivity for NTM detection warrants careful interpretation of these results. While high specificity (100%) ensures reliable positive results, some NTM infections may be missed. This limitation is inherent to the assay’s design, as the manufacturer specifies that it does not detect all known NTM species. Such limitation are typical of multiplex assays with predefined detection panels reflecting the wide diversity of NTM and the lack of universally conserved molecular targets [5,11]. In this validation study, the diversity of NTM species was constrained, potentially not representing the wider range encountered in routine clinical settings; this limitation is predominantly analytical, indicating predefined assay coverage rather than inherent diagnostic inadequacy. In clinical practice, negative NTM results should be interpreted alongside clinical and microbiological findings, with additional methods such as culture or line probe assays remaining crucial in suspected cases. In clinical terms, missed NTM detection may delay appropriate species-specific treatment, reinforcing the need for complementary diagnostic tests. These findings highlight the broader diagnostic challenge of comprehensive NTM detection and underscore the need for continued refinement of molecular panels to expand species coverage [21,22].

### Limitations

The limitations of this study include its reliance on well-characterized culture isolates rather than direct clinical specimens. While this approach is standard for initial analytical validation, it likely inflates estimates of diagnostic accuracy. Consequently, the reported performance represents an upper-bound analytical accuracy rather than true real-world clinical effectiveness. This distinction is critical for interpreting these findings and planning subsequent clinical validation studies. The impact of spectrum bias is particularly important for specific patient populations. For smear-negative and extrapulmonary tuberculosis, where a lower bacillary burden and the presence of inhibitors in clinical matrices can significantly reduce the sensitivity of molecular assays. We don’t know how well this assay will perform in these critical patient groups and requires dedicated clinical validation studies. Similarly, the impact of specimen type (e.g., sputum, cerebrospinal fluid, tissue, etc.), specimen quality, and storage conditions on assay performance has not been evaluated. Also, the use of a balanced sample distribution (1:1) may have influenced agreement statistics so the κ values should therefore be interpreted in the context of study design. Furthermore, the limited diversity of NTM species may not fully reflect real-world epidemiology. As with all molecular assays, detection is confined to predefined genetic targets, potentially missing rare or novel resistance-conferring mutations. Finally, operational factors like how well this assay works with different types of specimens, how much time it takes to do hands-on work, indeterminate rates, and cost-effectiveness, were not directly evaluated and need further study under programmatic settings.

### Pathway to Clinical validation

This analytical validation provides necessary foundation for clinical validations studies. The next crucial step is to evaluate assay performance on direct clinical specimen from patients with suspected TB including diverse specimen types and patient populations. Following successful clinical validation, implementation studies in reference laboratories and high throughput diagnostic centers will be essential to assess programmatic impact including effect on diagnostic turn-around-time, treatment initiation and patient outcome.

## Conclusion

This analytical validation demonstrates that multiplex open real-time PCR assays can achieve high analytical accuracy for simultaneous detection of MTBc, NTM differentiation, and identification of first-line drug resistance. The detection of isoniazid resistance represents a significant advance over existing rapid molecular platforms. The open-platform design offers important advantages for high-burden settings, including flexibility, cost optimization potential, and independence from proprietary supply chains.

However, the translation of these analytical findings to clinical practice requires systematic clinical validation on direct specimens, assessment of performance in diverse patient populations, and evaluation of programmatic feasibility and cost-effectiveness. When strategically deployed following clinical validation, these platforms may have the potential to enhance diagnostic precision, optimize treatment selection, and strengthen overall programmatic efficiency in TB control efforts.

## Data Availability

All data produced in the present work are contained in the manuscript

## Notes

**Competing interests:** All authors have completed the ICMJE uniform disclosure form and declare no support from any organization for the submitted work, no financial relationships with relevant organizations in the past three years, and no other relationships or activities that could have influenced the work.

### Competing Interest Statement

The authors have declared no competing interest.

### Funding Statement

This study did not receive any funding

### Author Declarations

The Ethics committee of New Delhi Tuberculosis Centre gave Ethical approval for this work

